# Equivalent SARS-CoV-2 viral loads between nasopharyngeal swab and saliva in symptomatic patients

**DOI:** 10.1101/2020.09.01.20186254

**Authors:** Isao Yokota, Takeshi Hattori, Peter Y Shane, Satoshi Konno, Atsushi Nagasaka, Kimihiro Takeyabu, Shinichi Fujisawa, Mutsumi Nishida, Takanori Teshima

**Author notes:** **Correspondence address** Takanori Teshima, M.D., Ph.D., Department of Hematology, Hokkaido University Faculty of Medicine, N15 W7, Kita-ku, Sapporo, Hokkaido 060-8638, Japan.

## Abstract

COVID-19 is diagnosed by detecting SARS-CoV-2 by nasopharyngeal swab (NPS) using real-time quantitative reverse transcriptase polymerase chain reaction (qRT-PCR). Emerging evidences have shown the utility of saliva, although conflicting results have been reported regarding viral loads between NPS and saliva. We conducted a study to compare the viral loads in 42 patients with COVID-19. Both NPS and saliva specimens were simultaneously obtained at a median of 6 days (range, 1-12) after symptom onset. SARS-CoV-2 was detected in 34 (81%) using NPS (median Ct value [IQR]=27.4 [21.3, 35.6]) and 38 (90%) using saliva (median Ct value [IQR]= 28.9 [23.1, 33.6]). There was no significance difference between them (Wilcoxon signed rank test: P=0.79) and Kendall’s *W* was 0.82, showing a high degree of agreement, indicating equivalent viral loads in NPS and saliva. After symptom onset, the Ct values of both NPS and saliva continued to increase over time, with no substantial difference. Self-collected saliva has a detection sensitivity comparable to that of NPS and is a useful diagnostic tool with mitigating uncomfortable process and the risk of aerosol transmission to healthcare workers.

## Introduction

Rapid and accurate diagnosis of coronavirus disease 2019 (COVID-19) is critical for containing outbreaks that may overwhelm healthcare systems. Although the detection of SARS-CoV-2 nucleic acids from nasopharyngeal swabs (NPS) is considered a gold standard in the diagnosis, self-collected saliva has been reported to have several advantages (1, 2). Specifically, self-collection reduces the risk of viral exposure for the healthcare worker and causes less discomfort for the patients. However, although emerging evidences have shown the utility of saliva as an alternative to NPS, (3-7) conflicting results have been reported regarding SARS-CoV-2 viral loads between NPS and saliva. Wyllie et al. showed that the viral load was five-times higher in saliva than NPS(8), while others have reported results to the contrary(9, 10). Herein, we conducted a study to compare the viral loads in paired samples (saliva and NPS) from symptomatic patients who were admitted for COVID-19.

## Methods

Forty-two patients diagnosed with COVID-19 by positive qRT-PCR of NPS were enrolled in this study. Paired NPS and saliva samples were simultaneously collected from all patients upon hospital admission between June 12, 2020 and August 6, 2020. This study was approved by the Institutional Ethics Board (Hokkaido University Hospital Division of Clinical Research Administration Number: 020-0116) and informed consent was obtained from all patients.

qRT-PCR was performed at a central laboratory (SRL, Tokyo, Japan). Self-collected saliva was diluted 4-fold with phosphate buffered saline and centrifuged at 2000 × g for 5 min to remove cells and debris. RNA was extracted from 200 μL of the supernatant or nasopharyngeal swab samples using QIAsymphony DSP Virus/Pathogen kit and QIAamp Viral RNA Mini Kit (QIAGEN, Hilden, Germany). qRT-PCR tests were performed, according to the manual by the National Institute of Infectious Diseases (NIID, https://www.niid.go.jp/niid/images/epi/corona/2019-nCoVmanual20200217-en.pdf). Briefly, 5uL of the extracted RNA was used to perform one step qRT-PCR using THUNDERBIRD® Probe One-step qRT-PCR Kit (TOYOBO, Osaka, Japan) and 7500 Real-time PCR Systems (Thermo Fisher Scientific, Waltham, USA). The cycle threshold (Ct)-values were obtained by using N1 primers (N_Sarbeco_F1, N_Sarbeco_R1) with N1 probe (N_Sarbeco_P1) and by using N2 primers (NIID_2019-nCOV_N_F2, N11 D_2019-nCOV_N_R2) with N2 probe (NIID_2019-nCOV_N_P2).

Ct values of qRT-PCR using NPS and saliva were expressed as scatter plots with Kendall’s coefficient of concordance *W* as nonparametric intraclass correlation coefficient. Scatter plots of Ct values and days from symptom onset for each type of specimen were also provided to examine the relationship between disease course and viral load. To find the longitudinal trends, we performed a median spline regression using “qsreg” function with the default parameters in R. Statistical analysis was conducted by R 4.0.2. All analyzed data were distributed in Supplement.

## Results

17 female (40%) and 25 male (60%) patients participated in the study. Median age of the patients was 73 years-old (range, 27 to 93) and specimens were obtained at a median of 6 days (range, 1-12) after symptom onset. SARS-CoV-2 was detected in NPS and saliva in 81% (34/42) and 90% (38/42) of the patients, respectively (Table 1). The Ct values using the N2 primers/probe were not significantly different between NPS and saliva, with median [IQR] of 27.4 [21.3, 35.6] and 28.9 [23.1, 33.6], respectively (Wilcoxon’s signed rank P=0.79, Figure 1A). Kendall’s *W* was 0.82, showing a high degree of agreement. Additionally, the Ct values of both NPS and saliva continued to increase over time, with no substantial difference (Figure 1C). There were cases of both NPS and saliva that became undetermined only three days after the onset of symptoms. Similar results were obtained from N1 primers/probe; the Ct values using the N1 primers/probe were equivalent between NPS and saliva, with median [IQR] of 31.0 [24.2, 39.5] and 33.1 [27.3, 37.3], respectively (Wilcoxon’s signed rank P=0.24, Figure 1B). Kendall’s *W* was 0.83, showing a high degree of agreement. Changes in Ct values of both NPS and saliva were not different over time (Figure 1D).

**Table 1.**
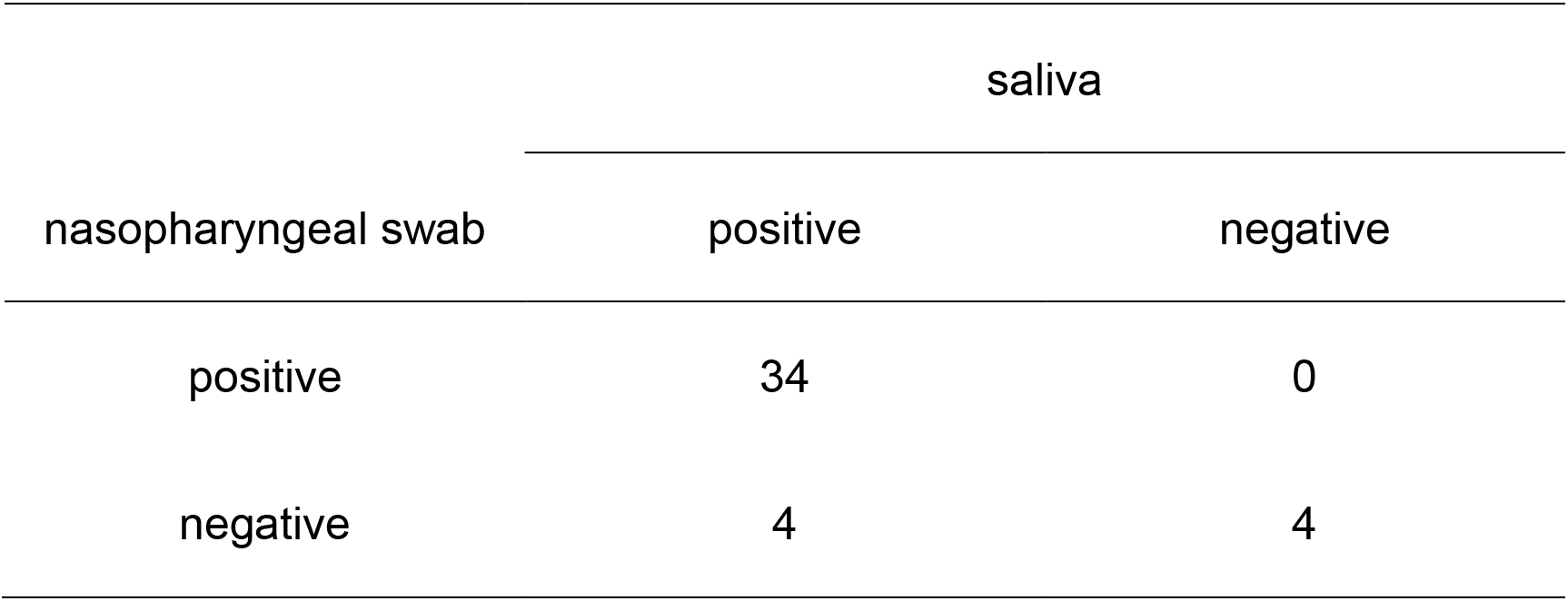
Detection summary of SARS-CoV-2 (N=42)

| nasopharyngeal swab | saliva |  |
| --- | --- | --- |
|  | positive | negative |
| positive | 34 | 0 |
| negative | 4 | 4 |

**Figure 1.**
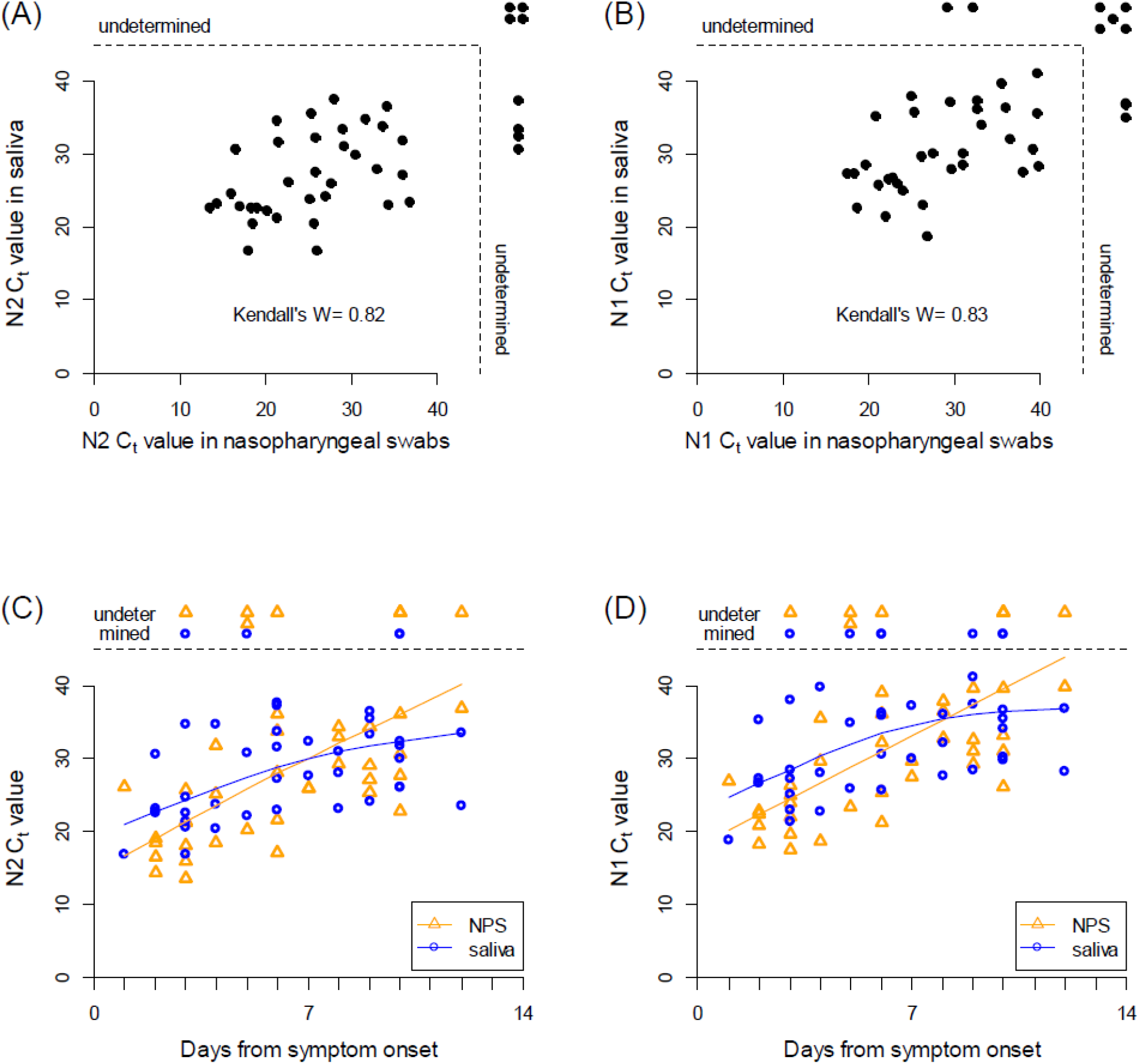
Viral loads of SARS-CoV-2 between nasopharyngeal swab and saliva specimens. **(A, B)** Scatter plots of Ct values using N2 **(A)** or N1 **(B)** primer and probe between NPS and saliva specimens taken from 42 COVID-19 patients. **(C, D)** Scatter plots of Ct values using N2 **(C)** or N1 **(D)** primer and probe primer against days from symptom onset. Median spline curves are also drawn using “qsreg” function with the default parameters in R.

## Discussion

In this study in symptomatic inpatients with COVID-19, SARS-CoV-2 was detected in saliva in 90% of the patients compared to 81% in NPS, with equivalent viral loads in the two specimens. Although there have been conflicting results reported to date(8-10), our study was designed to have significant advantages over previous studies; the number of patients was relatively large, paired samples were simultaneously collected, and qRT-PCR was performed at an independent central laboratory. Our results demonstrate that self-collected saliva is a useful alternative to NPS for the diagnosis of COVID-19. Furthermore, we recently reported equivalent sensitivity and specificity of qRT-PCR using saliva and NPS, again with equivalent viral loads in a large number of asymptomatic individuals in the setting of mass-screening(11). Taken together, self-collected saliva provides highly accurate results and should be considered as an easier and cost-efficient alternative for the detection of SARS-CoV-2 in both symptomatic and asymptomatic individuals.

In summary, self-collected saliva is a useful alternative to NPS as a specimen for detecting SARS-CoV-2 nucleic acids. The methodology of self-collection carries significant logistical and cost advantages over NPS by mitigating the risk of aerosol transmission to healthcare workers and obviating the need for full protective suits.

## Data Availability

N/A

## Acknowledgments

The authors declare that they have no competing interests. This study was funded by Health, Labour and Welfare Policy Research Grants 20HA2002.

## Notes

### Competing Interest Statement

The authors have declared no competing interest.

### Funding Statement

Yes described in the text

### Author Declarations

This study was approved by IRB

